# Cyclic Alternating Patterns (CAP) framework in Sleep Microstructure of Sleep Disorders: Markers of Sleep Instability Using Healthy Controls as Reference

**DOI:** 10.1101/2025.10.13.25337880

**Authors:** SI Dimitriadis, CI Salis

**Author notes:** **Correspondence to:** Dr. Stavros I. Dimitriadis, Head of Integrative Neuroimaging Lab, Thessaloniki, 55133, Makedonia, Greece.

## Abstract

This study introduces a novel multi-feature Sleep Instability Score (SLEIS) to assess sleep disorders. We evaluate its performance in distinguishing among seven sleep disorders, using a healthy control group as a reference. For the first time, our study extracts an exhaustive set of macrostructural and microstructural CAP sleep features from an open sleep disorder database. We measured the deviation from the healthy control group for all extracted features, quantifying effect sizes with Cohen’s *d*. We produced two versions of the SLEIS score: one where the individual feature value is multiplied by its corresponding Cohen’s d, and another based on cumulative weights over feature groups. A Random Forest (RF) model was used to rank the features that best distinguish the seven sleep disorders. This approach helped us identify a novel multi-feature marker of sleep instability. RF classification on the original feature values, using an eight-class approach, failed to robustly discriminate between disorders and healthy controls (precision = 56.44%, recall = 60%, F1-score = 57.87%). Both SLEIS versions led to clear improvement (feature groups/individual features: precision = 95.23% / 100%, recall = 90.71% / 100%, F1-score = 92.23% / 100%). Weighting macro- and microstructural features by their effect sizes, as deviations from a normative sample, is key. Our approach offers a promising solution for defining the new SLEIS marker that accounts for the heterogeneity of sleep disorders.

## 1. Introduction

The organization and progression of sleep stages at night are defined by cycles between non-rapid eye movement (NREM) and rapid eye movement (REM) sleep. This statement briefly describes sleep architecture (Mogavero et al. 2025). Each stage, including NREM substages (N1, N2, N3, and/or N4), is marked by distinct physiological processes, neural activity, and electroencephalographic (EEG) patterns (Troester 2023). Neurophysiologists describe NREM stages as periods of reduced arousal, relative quiet, and slower brain activity compared to wakefulness (Martin et al. 2020). In contrast, REM sleep is characterized by heightened brain activity, rapid eye movements, and vivid dreaming (Carskadon and Dement 2005).

The temporal organization of NREM-REM cycles shows the macrostructure of sleep. Several macrostructural parameters indicate timing, stage changes, and continuity, all from polysomnography (see Table 1 in Parrino et al. 2021). Sleep macrostructure refers to the overall arrangement of sleep stages across the night, including the time spent in each stage (N1, N2, N3, N4, REM) and the transitions between them. It gives the “big picture” of sleep architecture, highlighting major patterns and durations. Basic macrostructure measures are Total Sleep Time (total sleep duration), Sleep Latency (time to fall asleep), number of awakenings, NREM-REM duration, **Sleep Efficiency** (percentage of time in bed spent asleep), and Heart Rate (Ghermezian et al. 2023).

**Table 1.** Demographics of the sleep disorder dataset.

| Sleep disorder | No. of subjects | Males | Females |
| --- | --- | --- | --- |
| Bruxism (BRUX) | 2 | 2 (28.50 ± 7.77) | — |
| Sleep-disordered breathing (SBD) | 3 | 4 (71.25 ± 7.22) | — |
| Insomnia (INS) | 9 | 4 (68.00 ± 11.88) | 5 (55.20 ± 4.96) |
| Narcolepsy (NARCO) | 5 | 2 (33.50 ± 13.43) | 3 (30.33 ± 13.05) |
| Nocturnal frontal lobe epilepsy (NFLE) | 40 | 21 (27.80 ± 8.27) | 19 (33.00 ± 12.87) |
| Periodic leg movements (PLMs) | 10 | 7 (56.71 ± 7.54) | 3 (51.33 ± 1.15) |
| REM behavior disorder (RBD) | 22 | 19 (70.10 ± 6.65) | 3 (74.66 ± 1.52) |
| No pathology (N) | 16 | 7 (30.42 ± 3.73) | 9 (33.55 ± 6.52) |

In mammals, the master sleep clock regulates the harmonic changes of sleep and wake cycles over a 24-hour period. This clock is located in the suprachiasmatic nucleus (SCN) (Ma and Morrison 2025). The SCN is a small region in the hypothalamus. It regulates daily physiological and behavioral cycles, including the sleep-wake cycle (Moore 2007). The SCN receives light input from the eyes and synchronizes rhythms with the outside light-dark cycle. This process helps align our bodies with the 24-hour day (Mieda 2020). Many body functions exhibit oscillations with frequencies ranging from milliseconds to days. Examples include heart rate, brain activity, breathing, the sleep-wakefulness cycle, and the circadian rhythm. Circadian rhythms are oscillations with a roughly 24-hour period that are found in most physiological processes (Hastings et al. 2018). The SCN of the hypothalamus acts as a central circadian pacemaker in mammals. It syncs to the environmental light/dark cycle and coordinates circadian rhythms across many brain areas and tissues (Mieda 2020).

As aforementioned, the SCN regulates the harmonic alterations in sleep-wake cycles over a near 24-h period, and both environmental cues and genetic predisposition continuously modulate the SCN’s functionality (Jones et al. 2016). The contribution of environment, genes, and gene-environment interactions to sleep disorders is well established. Familial and twin sleep disorder studies revealed an important influence of genetic factors. Only four rare diseases may result from just a single mutation, and these are: familial advanced sleep-phase syndrome, fatal familial insomnia, chronic primary insomnia, and narcolepsy with cataplexy. These are called monogenic sleep disorders (Bidaki et al. 2012). However, most sleep disorders are more complex in their genetic susceptibility, including variation in the phenotype’s expressivity across family members (Dauvilliers and Tafti 2008).

### FROM SLEEP MACROSTRUCTURE TO CYCLIC ALTERNATING PATTERN

Sleep macro-architecture is an ordered sequence of altered sleep stages integrated within a sleep cycle. During sleep, mammals are not totally isolated from the external world. Rather, sleep allows interactions and integrations between the environment and internal habitats. Arousals are not open windows of wakefulness. Instead, they indicate the resistance of sleep macrostructure to potential perturbations. A single arousal does not impact an individual’s health or sleep quality. However, with physiological sleep, arousals may appear in swarms or as isolated events. Periodic and ordered sequences of arousals form the so-called cyclic alternating pattern (CAP), as defined by Terzano (Terzano 1991).

Terzano et al. introduced CAP to sleep research based on their observation that microstructural events occur in bursts rather than in isolation (Terzano et al. 1985). They divided sleep into two phases: A and B. Phase A involves phasic events that deviate from the EEG background activity. It includes K-complex sequences, delta bursts, vertex sharp transients, arousals, and polyphasic bursts. Phase B represents tonic, non-transient background activity. Phase A can be subdivided into three subtypes (see Materials and Methods). Sleep at the microstructural level is classified as CAP if a phase A is followed by a phase B within 60 seconds. It is classified as non-CAP if only phase B is present. The most frequent statistic extracted from CAP sleep microstructure is the CAP rate. This is estimated as the percentage ratio of total CAP time to total NREM sleep time (CAP + non-CAP). A heightened presence of phase A is seen as sleep disturbances at the microstructural level. The CAP rate is considered a measure of arousal instability. Higher CAP rates are associated with poorer sleep quality (Parrino et al. 2012).

Altered CAP rates have been found in many pathological sleep conditions, including sleep apnea, insomnia, and periodic limb movements (Parrino et al. 1996, 2012; Terzano et al. 1996; Zucconi et al. 2000). Disorders such as bruxism, sleepwalking, and epileptic activity are more common during phase A, while sleep apneas usually occur during phase B (Parrino et al. 2012).

The present study aims to quantify CAP-related microstructural features in a healthy control group and seven sleep disorders. This may reveal potential markers of sleep instability. We analyzed 108 polysomnographic recordings from the CAP Sleep Database, contributed by the Sleep Disorders Center of the Ospedale Maggiore of Parma, Italy (Goldberger et al. 2000; Terzano et al. 2001). Our microstructural analysis focused on annotated CAP over the hypnograms provided by Terzano’s group. We also estimated macrostructural features from hypnograms to explore their complementarity with microstructural CAP feature sets. Recent neuroimaging studies have used normative modeling to detect brain disorders as deviations from age-matched charts across multiple modalities. Similarly, we use the healthy control group in this database as a reference for the seven sleep disorders. We anticipate that robust, sleep-disorder prior effect size weights, referenced to a common healthy control group, will help parse individual-level and disorder-specific heterogeneity. This approach accounts for personalized profiles of macrostructural and microstructural alterations. It is expected to provide sleep disorder–specific sensitivity.

The remainder of this paper is organized as follows. Section 2 describes the existing CAP database and the quantified macrostructural and microstructural CAP features. Section 3 presents the results. Section 4 discusses the added value, limitations, and future directions of this work.

## 2. Materials and Methods

### 2.1. Material

For the purpose of our study, we downloaded all-night polysomnography (PSG) recordings from the well-known database: “The CAP Sleep Database” of “PhysioBank” (https://physionet.org/physiobank/database/capslpdb/) (Goldberger et al. 2000; Terzano et al. 2001). This study comprises 108 PSG recordings from seven groups with sleep disorders and a normal healthy group that did not present any medical, neurological, or psychiatric disorders. The study has been approved by the Sleep Disorders Center of the Ospedale Maggiore of Parma, Italy (Goldberger et al. 2000; Terzano et al. 2001). The clinical information for the entire cohort is available in the Excel file “gender-age.xlsx”. Sleep recordings have been manually annotated by well-trained sleep expert neurologists, according to the Rechtschaffen & Kales rules, classifying epochs into sleep stages 1–4, wake, REM sleep, and movement artifacts (Wolpert 1969). Cycling Alternating Pattern (CAP) was determined based upon Terzano’s rules (see the following section) (Terzano et al. 2001).

**Table 1** summarizes the number of subjects per sleep disorder, with their gender distribution and mean age. The number of participants in our study with the related sleep disorder is provided in **Table 2**.

**Table 2.** A summarization of the indices of the selected subjects per group.

| Bruxism | Insomnia | Healthy controls | Narcolepsy | Nocturnal frontal lobe epilepsy | Periodic leg movements | REM behavior disorder | Sleep-disordered breathing |
| --- | --- | --- | --- | --- | --- | --- | --- |
| Brux1, brux2 | isn1-ins9 | n1-n5, n10, n11, n16 | Narco1-narco5 | nfle1-nfle40 | plm1-plm10 | rbd1-rbd22 | sdb1-sdb4 |

### 2.2 Methodological and technical requirements for scoring a CAP sequence

An important processing step before CAP identification is the manual sleep-stage annotation by sleep experts, based on conventional criteria (Wolpert 1969; Iber 2007).

#### 2.2.1 Onset and termination of a CAP sequence

The CAP is a periodic EEG pattern that occurs during NREM sleep. A CAP is characterized by cyclic sequences that are composed of a succession of CAP cycles. A CAP cycle is composed of a phase A followed by a phase B, and at least two CAP cycles are essential to form a CAP sequence. All CAP sequences begin with a phase A, which underlines cerebral activation, and end with a phase B. Phase B comprises periods of deactivation that are interspersed by two successive phase A periods with intervals shorter than one minute. The duration of each CAP phase ranges from 2 to 60 seconds. This cut-off threshold has been defined based on the observation that more than 90% of A phases that occur during NREM sleep are separated by an interval of less than 1 minute (60 seconds) (Terzano et al. 1985; Terzano 1991; Parrino et al. 2012).

CAP is the EEG translation of the reorganization of the sleeping brain that is challenged by modifications in environmental conditions. Phases A and B are the basic compounds of each CAP cycle, corresponding to levels of greater and lesser arousal, respectively (Terzano et al. 1985). When CAP appears, arousal fluctuates constantly between two distinct sleep levels. This modulation of arousal affects not only the EEG but also the autonomic, muscular, and behavioral functions, which increase during phase A and decrease during phase B (Terzano et al. 1982; Parrino et al. 2012).

CAP expresses a basic arousal modulator that survives in conditions of severely impaired vigilance but essentially belongs to physiological normal sleep (Steriade et al. 1994). According to current findings, CAP appears to be a well-defined marker of cerebral activity under conditions of reduced vigilance (sleep, coma), reflecting a state of instability involving muscle, behavioral, and autonomic functions. The absence of CAP coincides (see following sub-section) with a condition of sustained arousal stability and is defined as non-CAP.

#### 2.2.2 Non-CAP

1. Below, we summarize the three rules for characterizing a period of sleep activity as a non-CAP.
2. A period without CAP lasting over 60 seconds is scored as non-CAP, representing stationary activity without any phasic phase A, regardless of NREM sleep stage.
3. An isolated phase A, separated from another phase A by more than 60 seconds, is counted as **non-CAP**.
4. A phase A that ends a CAP sequence is classified as **non-CAP**.

#### 2.2.3 Minimal criteria for the detection of a CAP sequence

There is no upper limit to the overall duration of the CAP sequence or the total number of CAP cycles. In young adults, a typical CAP sequence lasts about 2 minutes and 30 seconds and includes around 6 CAP cycles (Smerieri et al. 2007). As previously noted, a CAP sequence is defined by at least two consecutive CAP cycles (**Fig. 1**). To be considered a CAP sequence, three or more consecutive A phases must be observed, with each of the first two A phases followed by a B phase within 60 seconds, and the last A phase followed by a non-CAP interval longer than 60 seconds.

**Fig.1.**
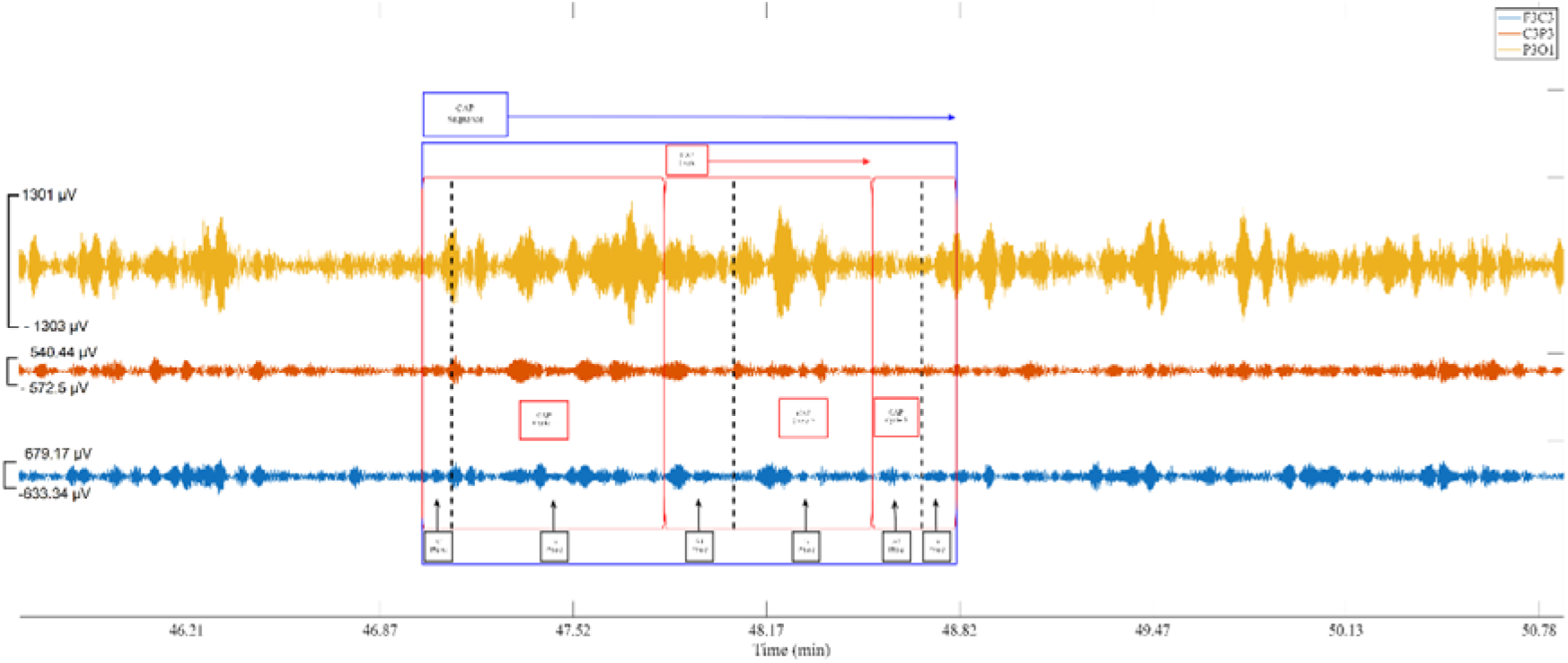
An example of a CAP sequence with the enclosed CAP cycles and CAP phases A and B from the first normal subject 1 (n1) extracted from the N1 sleep stage. The EEG signal was filtered within the 1-12 Hz frequency range using the eegfilt function and FIR filtering.

#### 2.2.4 General rules

Below, we summarize the general rules for defining a CAP sequence (Parrino et al., 2012).

1. A phase A counts in a CAP sequence only if it occurs within 2 to 60 seconds before or after another phase A.
2. A CAP sequence starts after a non-CAP NREM sleep EEG period over 60 seconds, except for three cases below.

There are no time limits:

before the first CAP sequence in non-REM sleep;
after transitioning from wake to sleep;
after moving from REM to non-REM sleep.

#### 2.2.5 Stage shifts

Within the NREM sleep, the evolution of a CAP sequence is not interrupted by a sleep stage shift if all CAP scoring rules are satisfied. For that reason, a CAP sequence can encapsulate different phase A and B activities, since it can extend across adjacent NREM sleep stages (Terzano 1990).

### 2.3 Types of A-Phases

A-phases can be divided in three types: (a) A1-phase is characterized by bursts of *K*-complexes or delta waves (0.5–4 Hz), (b) A2-phase has rapid (frequencies in the beta or alpha band) EEG waves that occur for 20–50 % of the total A-phase time, and (c) A3-phase is characterized by rapid waves, alpha (8–12 Hz) and beta (16–30 Hz), that occupy more than 50 % of the total A-phase time (see **Table 3**). Thus, each type of A-phase has distinct EEG frequency characteristics and different roles and distributions within the sleep cycle. In normal sleep, each A-phase type is related to the subject’s age, sleep stage, and time of sleep, whereas in pathological conditions, the A3-phase reflects alterations in sleep induced by noxious events, i.e., obstructive apnea and leg movement (Mendez et al. 2016).

**Table 3:**
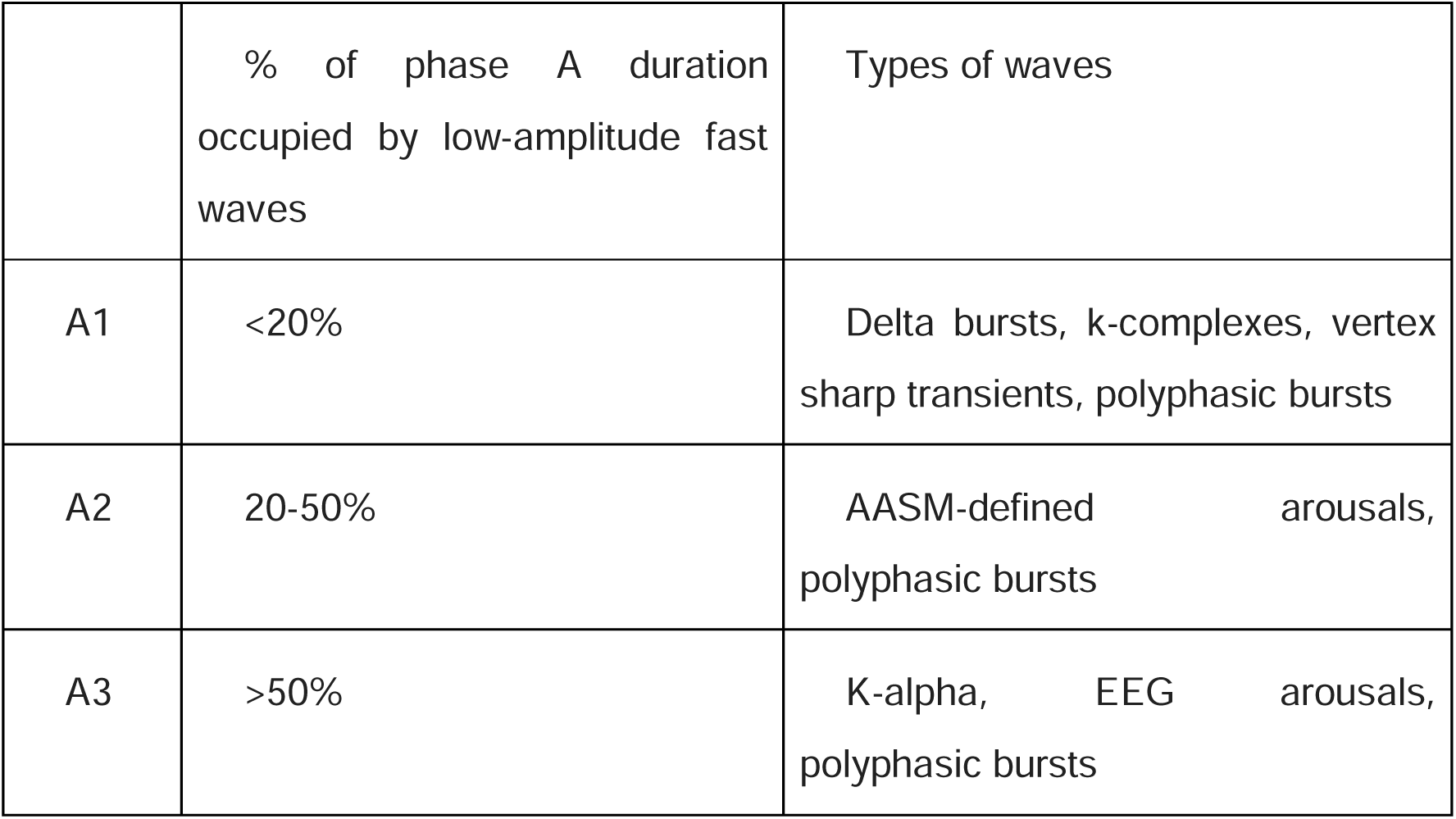
EEG characteristics of the different A phases in CAP.

|  | % of phase A duration occupied by low-amplitude fast waves | Types of waves |
| --- | --- | --- |
| A1 | <20% | Delta bursts, k-complexes, vertex sharp transients, polyphasic bursts |
| A2 | 20-50% | AASM-defined arousals, polyphasic bursts |
| A3 | >50% | K-alpha, EEG arousals, polyphasic bursts |

Physiologically, CAP A1 subtypes prevail in the descending branches of the first sleep cycles and gradually decrease during the night. This trend mirrors the decline of homeostasis. Conversely, CAP A2 and A3 dominate the ascending branches of sleep cycles and can be considered the forerunners of REM sleep (Terzano et al. 2005).

### 2.4 Macrostructure and Microstructure of Sleep

The macrostructure of sleep refers to the overall organization and timing of sleep stages throughout the night. It focuses on the sequence and duration of different sleep stages, including NREM (non-rapid eye movement) sleep (stages N1, N2, N3) and REM (rapid eye movement) sleep, and how these stages cycle through the night. CAP indicates a condition of physiological instability, while non-CAP indicates a condition of physiological stability. Non-CAP and CAP sequences constitute the microstructure of sleep. **Fig. 2** illustrates an example of each sleep disorder, including the hypnogram and the appearance of CAP sequences across the sleep stages.

**Fig.2.**
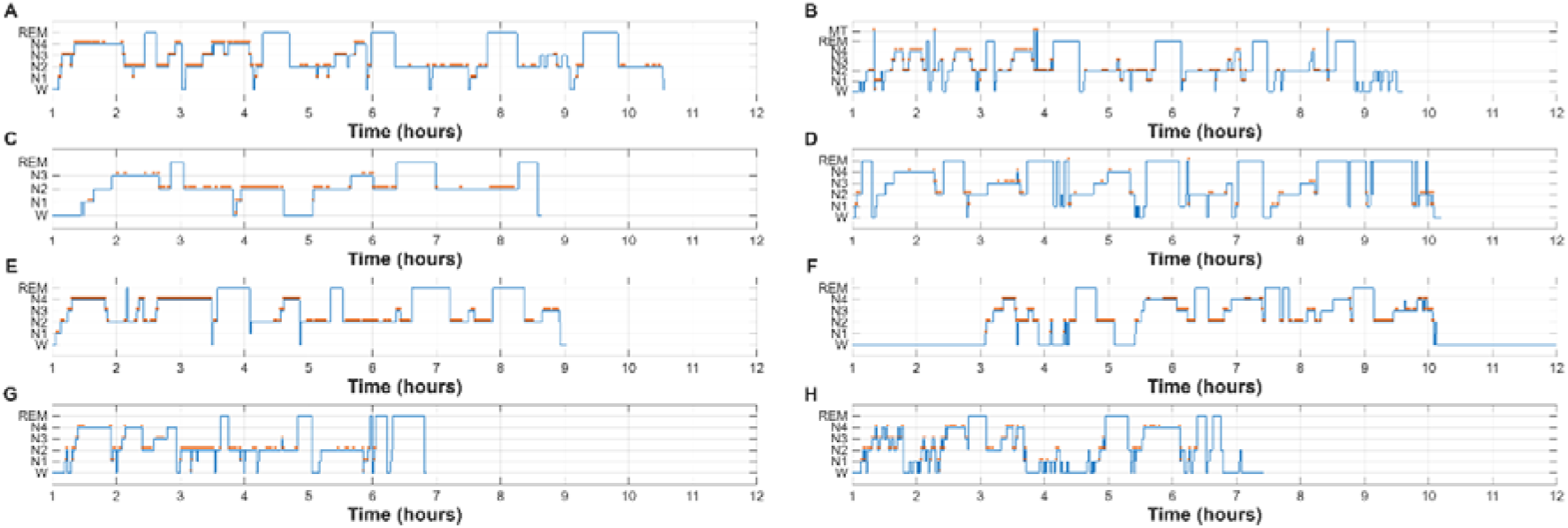
An example of hypnograms and the appearance of CAP sequences for each subject group and class is shown as red dots. A - subject n1, B - subject brux1, C - subject ins1, D - subject narco1 E - subject nfle1, F - subject plm1, G - subject rbd1, H - subject sdb1.

### 2.5 Description of Quantitative Features Extracted based on Hypnograms and CAP

We estimated a total number of one hundred and thirteen (113) microstructural and macrostructural features. Our analysis focused on the formation of CAP sequences for the microstructural extraction of CAP-related features.

#### 2.5.1 Macrostructural Sleep Features

This section includes the description of the eleven (11) macrostructure features.

##### Sleep Efficiency (%)

This feature is the ratio of the total time a person spends asleep to the total duration of sleep (including the wake state). Mathematically, Speep Efficiency can be expressed via the following equation:

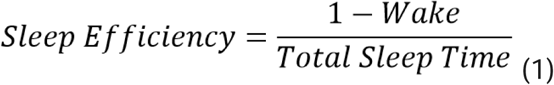

where indicates the total time the person is awake. This characteristic corresponds to 1 feature.

##### Ratio of Sleep Stage X to the total duration of NREM plus REM (%)

This characteristic measures the percentage of time that the brain spends in a specified stage, referred to as stage X, relative to the total time in NREM plus REM stages. Here, stage X represents any of the following: time spent in N1, N2, N3, or N4; the combined time in N1 and N2; the combined time in N3 and N4; total time in NREM; or total time in REM. As a result, this feature yields eight (8) features. Mathematically, this ratio is calculated as follows:

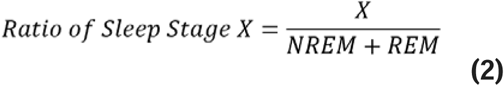

##### Total sleep time (minutes)

This characteristic measures total sleep time, i.e., the sum of NREM and REM stages, yielding one (1) feature.

##### Time passed from the first N1 epoch to the first REM epoch (minutes)

This feature measures the time passed from the beginning of the first N1 epoch until the first REM epoch. As a result, this quantity yields one (1) feature.

#### 2.5.2 Microstructural CAP features

This section describes the microstructural features. These characteristics are quantified within three distinct domains: e.g.

**Domain 1** - Whole NREM sleep: Encompassing stages N1, N2, N3, and N4

**Domain 2** – Light sleep: Encompassing stages N1 and N2

**Domain 3** – Deep sleep: Encompassing stages N3 and N4
while in some of them we adopted both the mean and the median as statistical metrics. This approach leads to a total of one hundred and two (102) microstructural features. In Domains 2 and 3, our analysis focused on CAP sequences that start and end within the same sleep stage (either N1 or N2, or N3 or N4). **Table 4** illustrates the definitions of the extracted features, divided into one measurement (estimation of mean/median in NREM) and mean/median in N1/N2 and N3/N4.

**Table 4.**
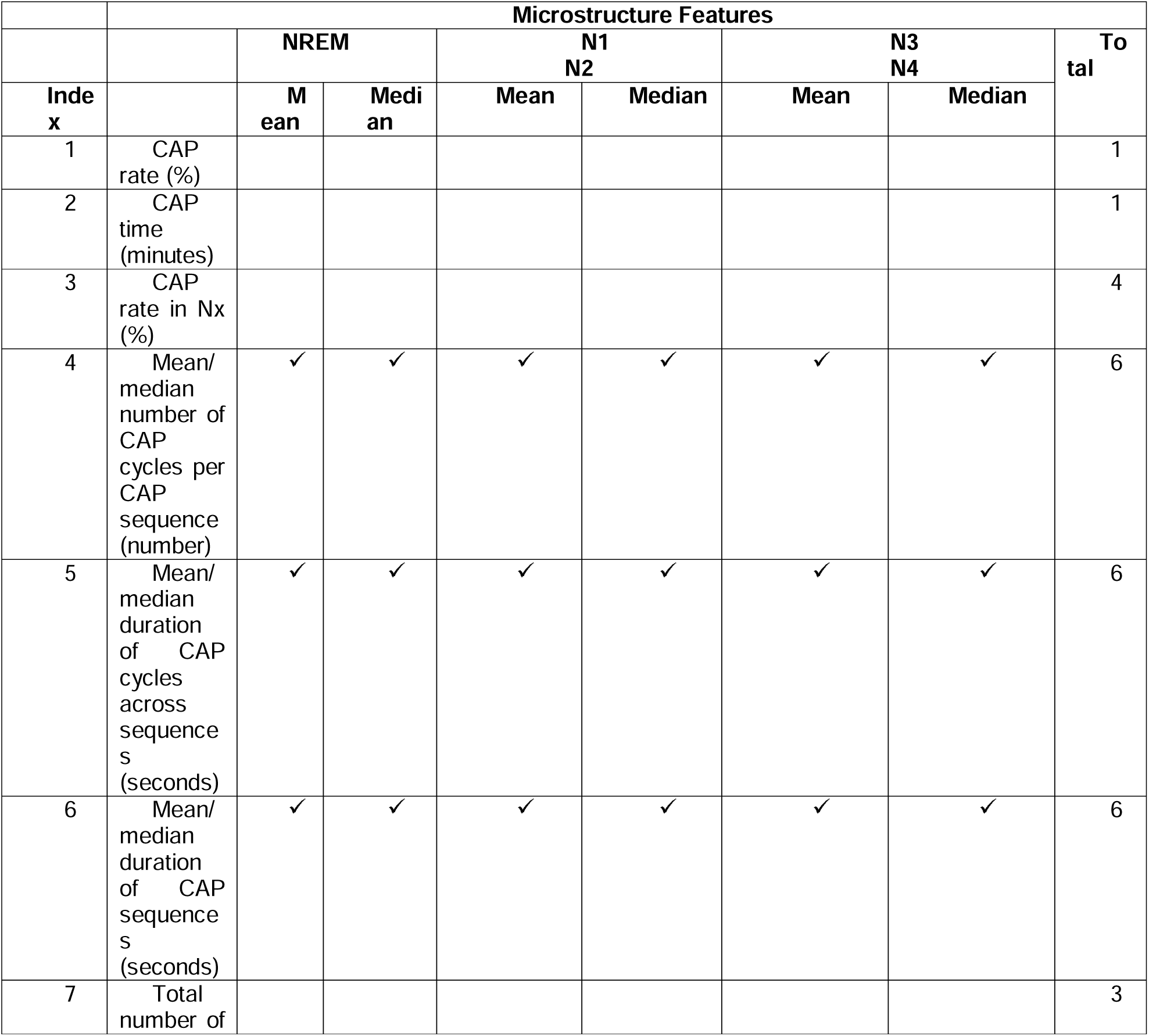
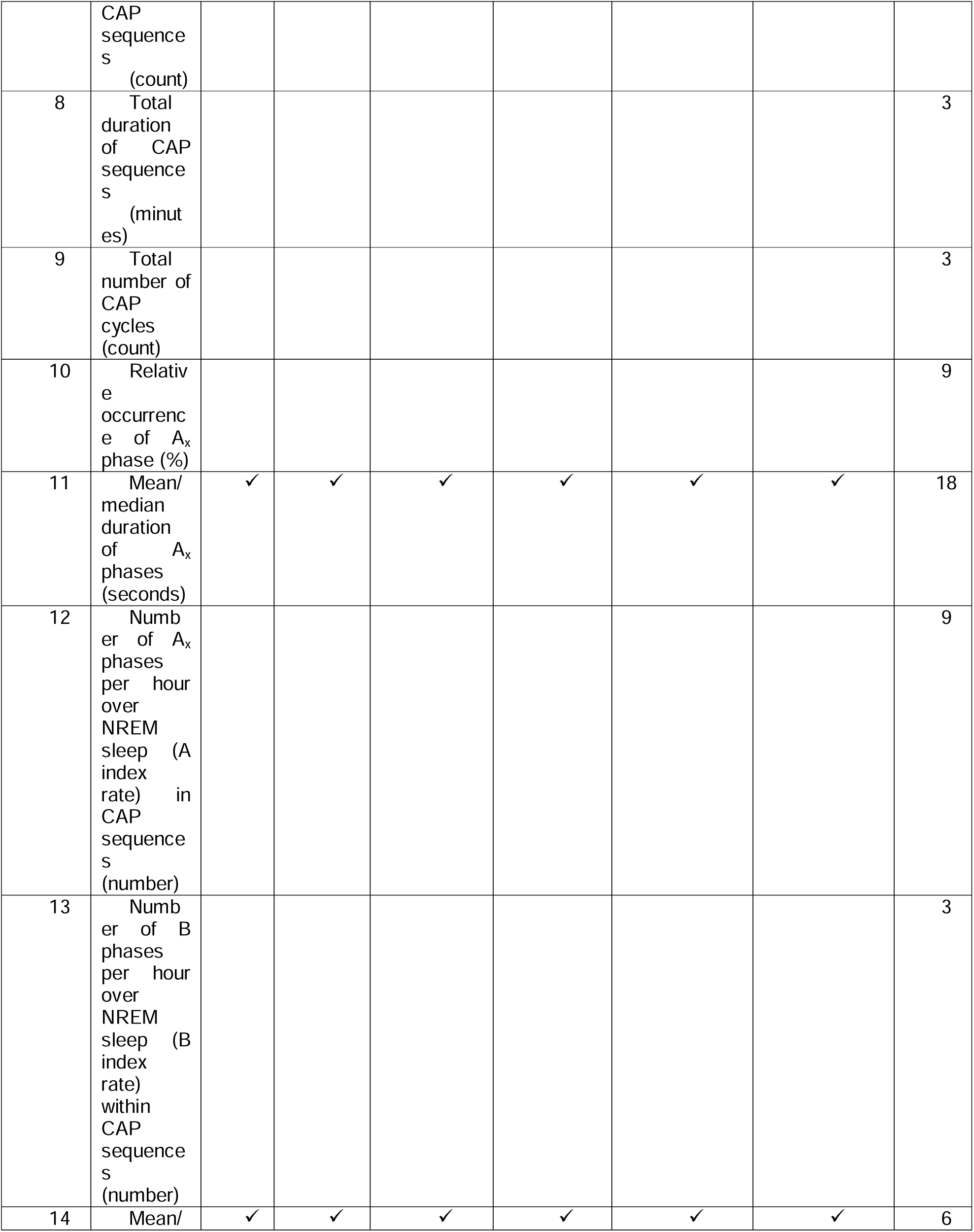
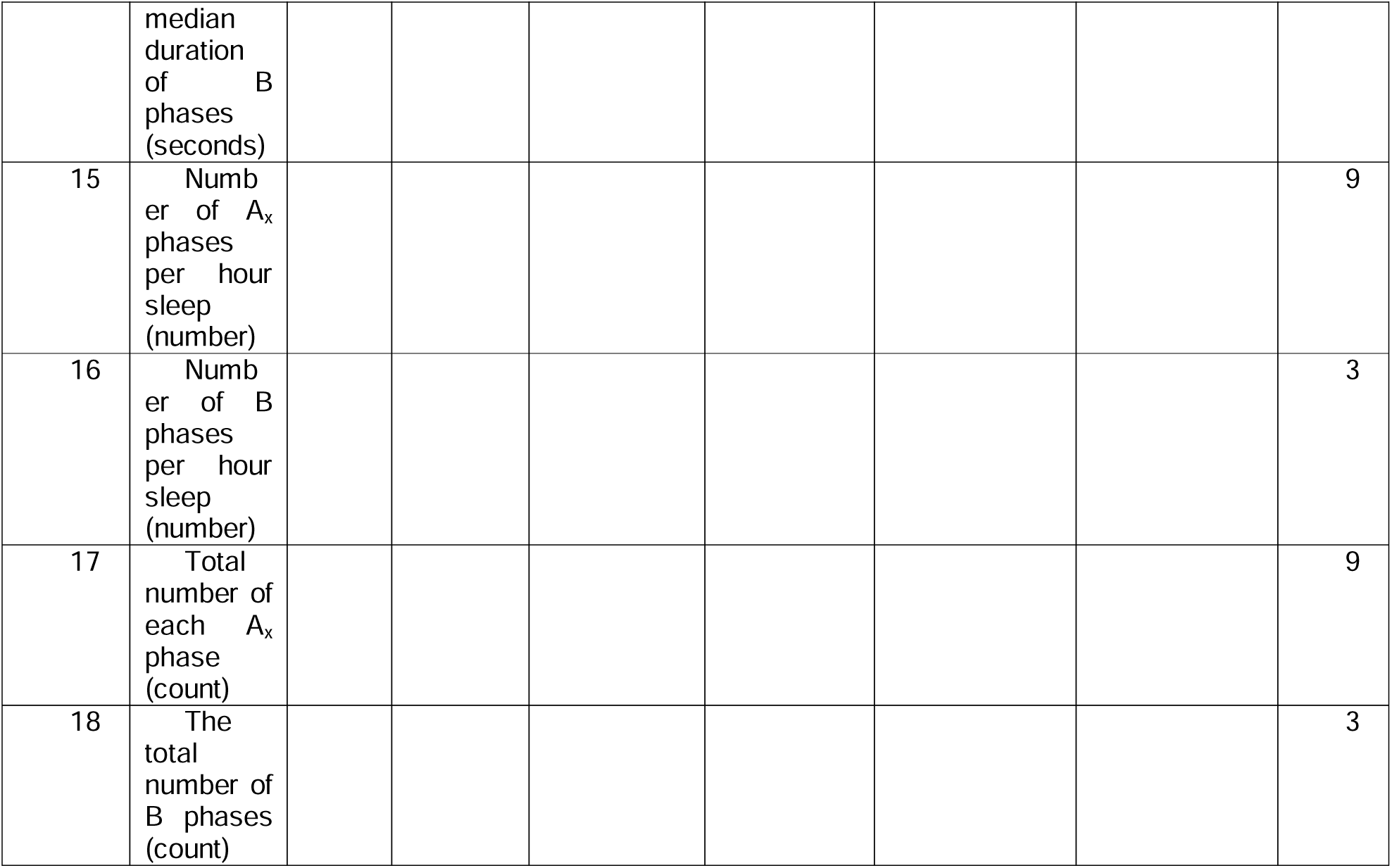
Definition of microstructure features across three domains (NREM, light (N1/N2), and deep (N3/N4) sleep) and in both approaches using mean and median. Features with no tick are estimated across the sleep cycle.

##### CAP rate (%)

This feature is defined as the ratio of the total time (in minutes) spent in all CAP sequences to the total time (in minutes) spent in NREM sleep. It is computed within **Domain 1** and yields one feature.

##### CAP time (minutes)

This feature is defined as the cumulative duration (in minutes) spent in all CAP sequences across NREM stages. It is computed within **Domain 1** and yields one feature.

##### CAP rate in N_x-_ (%)

This feature is defined as the duration spent in all CAP sequences related to stage Nx (where Nx can be N1, N2, N3, or N4) divided by the total duration of stage Nx. It is computed within **Domain 1**, resulting in four features (one for each stage).

##### Mean/median number of CAP cycles per CAP sequence (number)

This feature is computed by first counting the number of CAP cycles in each CAP sequence, then calculating the mean and median of the resulting distribution. As a result, this characteristic corresponds to two (2) sub-features (mean value and median) and is extracted for each of the **Domains 1, 2, and 3**, yielding a total of six (2×3) features.

##### Mean/median duration of CAP cycles across sequences (seconds)

This feature is computed by measuring the mean duration of all CAP cycles within each CAP sequence, then calculating both the mean and the median of the resulting distribution. As a result, this characteristic corresponds to two (2) sub-features (mean value and median) and is extracted for each of the **Domains 1, 2, and 3**, yielding a total of six (2×3) features.

##### Mean/median duration of CAP sequences (seconds)

This feature is computed by measuring the duration of each CAP sequence, then calculating the mean and median of the resulting distribution. As a result, this characteristic corresponds to two (2) features (mean value and median) and is extracted for each of the **Domains 1, 2, and 3**, yielding a total of six (2×3) features.

##### Total number of CAP sequences (count)

This feature is computed by measuring the number of CAP sequences across **Domains 1, 2, and 3, generating three (1×3) features**.

##### Total duration of CAP sequences (minutes)

This feature is computed by summing the durations of CAP sequences across **Domains 1, 2, and 3, generating three (1×3) features**.

##### The total number of CAP cycles (count)

This feature is computed by measuring the total number of CAP cycles across **Domains 1, 2, and 3, generating three (1×3) features**.

##### Relative occurrence of A_x_ phase (%)

This feature is defined as the number of Ax phases (where Ax corresponds to A1, A2, or A3 divided by the total sum of A1 + A2 + A3 phases. As a result, this feature yields three (3) sub-features (one for each Ax phase) and is computed across **Domains 1, 2, and 3**, resulting in a total of nine (3×3) sub-features.

##### Mean/median duration of A_x_ phases (seconds)

This feature is defined as the mean and the median duration of each Ax phase. The aforementioned quantity is calculated across **Domains 1, 2, and 3**, yielding two sub-features (mean and median) for each Ax phase per domain, and a total of eighteen (3×6) sub-features

##### Number of A_x_ phases per hour over NREM sleep (A index rate) in CAP sequences (number)

This characteristic is computed by dividing the number of Ax phases occurring within CAP sequences by the total NREM time (in seconds), then normalizing to hours by NREM time ÷ 3600. This process yields three sub-features (one for each Ax phase) and repeats across **Domains 1, 2, and 3**, resulting in nine (3×3) features.

##### Number of B phases per hour over NREM sleep (B index rate) within CAP sequences (number)

This characteristic is computed by dividing the total number of B phases within CAP sequences by the total NREM time (in seconds), then normalizing to hours (NREMtime ÷ 3600). The aforementioned quantity is computed across **Domains 1, 2, and 3**, yielding three (3) features.

##### Mean/median duration of B phases (seconds)

This feature is the mean and median durations (seconds) of B phases, calculated for **Domains 1, 2, and 3**, yielding two sub-features per domain and six features overall (2×3).

##### Number of A_x_ phases per hour (number)

This feature is the number of Ax phases in CAP and non-CAP sequences, divided by NREM time (hours), yielding one sub-feature per Ax phase per domain across **Domains 1, 2, and 3**, for a total of nine features (3×3).

##### Number of B phases per hour over NREM sleep (number)

This feature is the number of B phases in both CAP sequences and non-CAP epochs, divided by total NREM time (in hours, NREMtime ÷ 3600). It is computed for **Domains 1, 2, and 3**, resulting in three features.

##### Total number of A_x_ phases (count)

This feature is the total number of Ax phases in CAP and non-CAP epochs. One sub-feature is defined for each Ax phase across **Domains 1, 2, and 3**, yielding 9 features (3×3).

##### The total number of B phases (count)

This feature is the number of B phases in both CAP sequences and non-CAP epochs, calculated for Domains 1, 2, and 3, yielding three features.

### 2.6 Sleep Instability score (SLEIS) analysis

For each sleep disorder group, we calculated Cohen’s *d* effect sizes for each feature relative to the healthy control reference group. Each participant’s original feature value is then weighted by the absolute value of Cohen’s *d* effect size established by the comparison of the healthy control group with the relevant sleep disorder group. The following equation (Eq. 3) describes the aforementioned definition at the feature level.

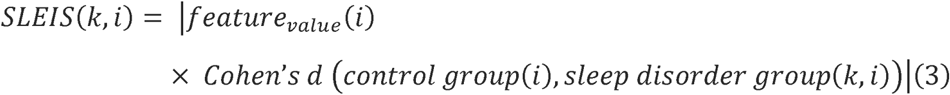

where (i) runs across the total number of features and (k) across the seven sleep disorder groups, and |.| indicates the corresponding absolute value.

Alternatively, we considered the aggregated Cohen’s *d* effect sizes for the four feature groups, as presented in section 2.5. The following equation (Eq.4)

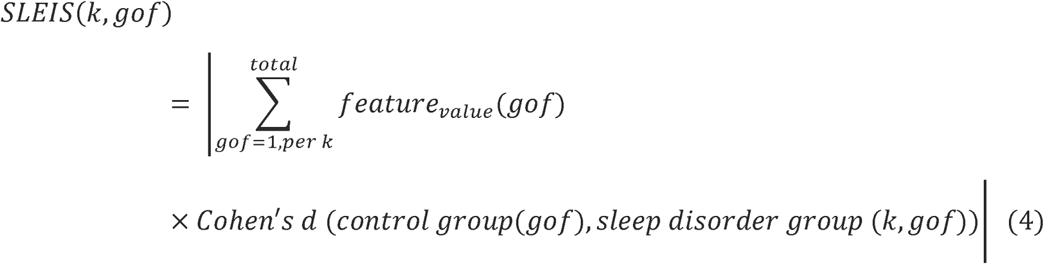

where (k) runs across the seven sleep disorder groups, gof (group of features) runs across the ***total*** number of each distinct feature set, and |.| indicates the corresponding absolute value. In both approaches, the absolute SLEIS score was taken.

This approach addresses the limited number of subjects in many sleep disorder groups while quantifying individual participant deviation from a within-sample norm tailored to the relevant sleep disorder group.

### 2.7 Random Forest (RF) model and experimental setup

#### 2.7.1 RF Classifier

The RF classifier is an ensemble learning method, used extensively in classification and regression tasks (Breiman 2001). The methodology essentially involves constructing a group (called a ‘forest’) of decision trees as follows: each tree in the ‘forest’ is built on a different training set (called a bootstrap sample), which is randomly drawn from the original training data. A randomly selected variable subset (from the original variable set) is used (during the construction of each decision tree) to establish the best split at each node. Once the RF model has been trained and all trees in the forest have been constructed, their decisions are combined using majority voting (for classification) or averaging (for regression). Then, these decisions are used in predicting unknown cases.

The RF algorithm provides an internal estimator of the generalization error of the constructed model, called out-of-bag (OOB) error. The estimation of the OOB error involves each decision tree in the model as follows: a portion of the original data cases (around 1/3) does not participate in the training of each tree and instead is used as ‘test’ data, being predicted by the constructed tree. The OOB error estimate is the average prediction error across all training cases, computed using only the trees that do not include that case in their bootstrap samples. For more details on the RF algorithm and its underlying notions, we refer, for example, to (Liparas et al. 2014; Dimitriadis et al. 2018a).

#### 2.7.2 Experimental setup

In all RF models trained in our experiments, the following parameter setup was used: the number of trees per model was empirically set based on the OOB error estimate. Following this approach, the number of trees was set to 1000 in all cases. For each RF model and each node split during tree growth, the number of variables used to determine the best split was set to the default √m (m = the total number of features in the dataset). In addition, the RF classifier has its own built-in feature selection method that selects the most important features for each RF model. Consequently, only the selected feature subsets were used to train the final RF models. Finally, a 10-fold cross-validation scheme was used in all experiments to train the classifier. It is worth mentioning that the code for our RF-related experiments was written in R, version 4.3.3 (www.r-project.org/).

#### 2.7.3 Experiments

Below, we summarize the experiments using the RF model and different combinations and definitions of our feature set.

##### Experiment A

We adopted the same approach as in our previous study, investigating how cross-frequency coupling is altered across sleep disorders in non-CAP epochs (Dimitriadis et al., 2021). Specifically, we used an eight-class classification problem (healthy control group plus seven sleep disorders) and the full feature set. This approach will serve as the reference model for the proposed approaches described in section 2.6 and in Experiments B and C.

##### Experiment B

We adopted the RF model with the four SLEIS scores corresponding to the four feature sets (eq. 2) for a seven-class classification problem (seven sleep disorders). Here, we grouped the features in four sets: A) macrostructural features (11), B) CAP rates (6), C) CAP cycles and sequences (27), and D) CAP Phases A and B (69).

##### Experiment C

We adopted the RF model with SLEIS scores for each feature (eq. 1) in a seven-class classification problem (seven sleep disorders).

The outcome of these experiments will quantify the importance of our approach and reveal the significance of SLEIS scores at both single-feature and feature-set levels.

### 2.8 Software

The code for analyzing annotated hypnograms with CAP-related information has been written in MATLAB (MathWorks, version 2024b). RF-related experiments have been conducted in R, version 4.3.3 (www.r-project.org/). All figures were generated in MATLAB and R.

## 3. Results

Before estimating the proposed SLEIS scores, eight features (out of 113) were excluded from the sample because they had zero values in a small number of cases. A small number of subjects had zero values in this set of features, corresponding to sleep disorder groups with few samples. A small number of subjects had zero values in this set of features, corresponding to sleep disorder groups with few samples. As a result, the estimation of SLEIS scores was conducted with one hundred and five (105) features.

### 3.1 Performance of the RF model with eight classes

Our RF model achieved high performance of 87.96% in the eight-class scenario, but with macro-average precision of 56.44%, macro-average recall of 60%, and macro-average *F*-score of 57.87% (**Table 5**). It is important to note that the RF model achieved the highest classification accuracy for the normal, insomnia, and NFL groups; however, no samples were classified as SDB. The OOB estimate of error rate was 51.69%. **Table 6** summarizes the confusion matrix of the RF model for the eight-class approach. The seven most discriminative features, based on our RF model, with a Gini Index> 1 and a z-score > 1.5 across all features, are shown in **Fig. 3**.

**Fig.3.**
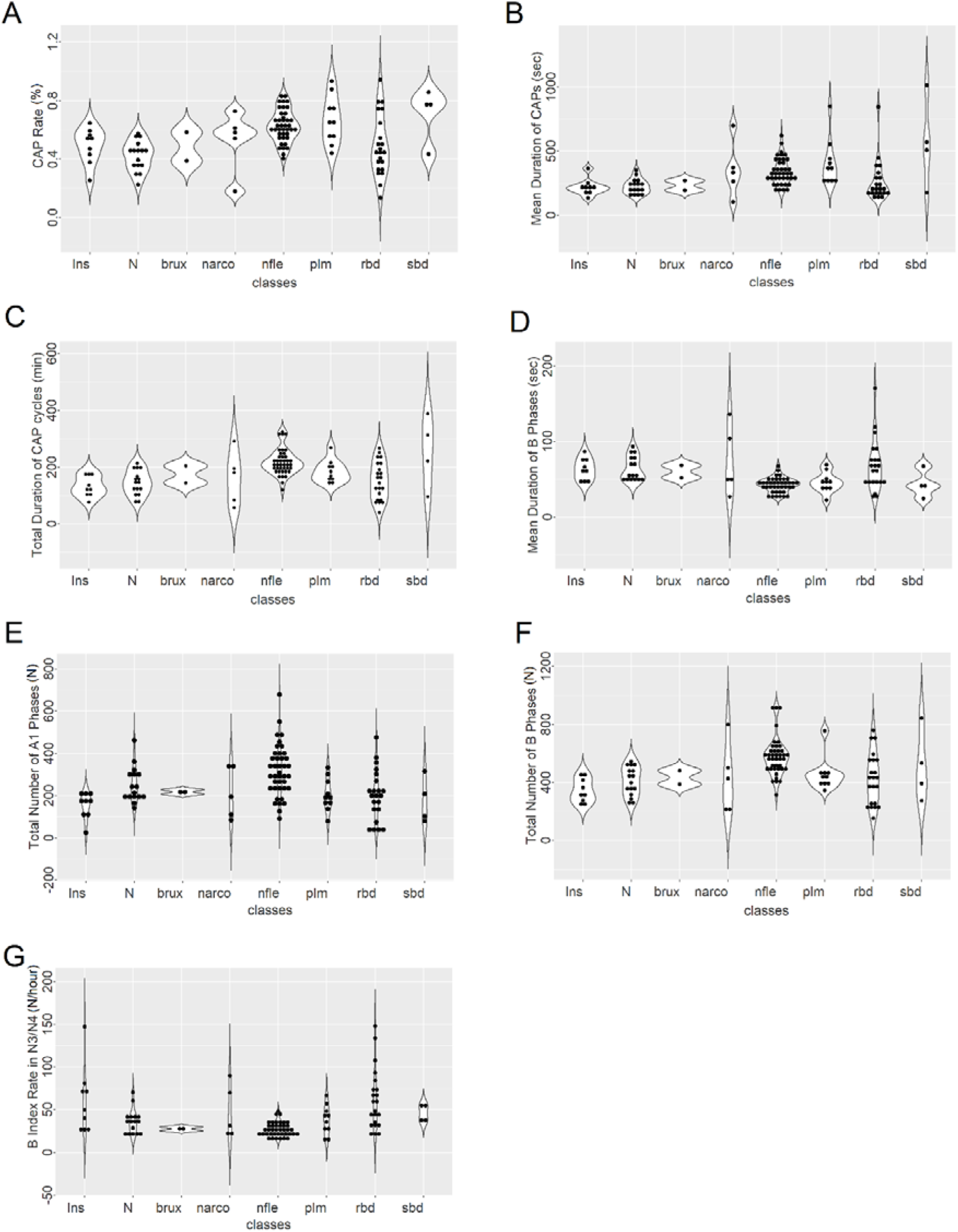
Mean and standard deviations of the seven most discriminative features across the normal subject group and the seven sleep disorder groups.

**Table 5.**
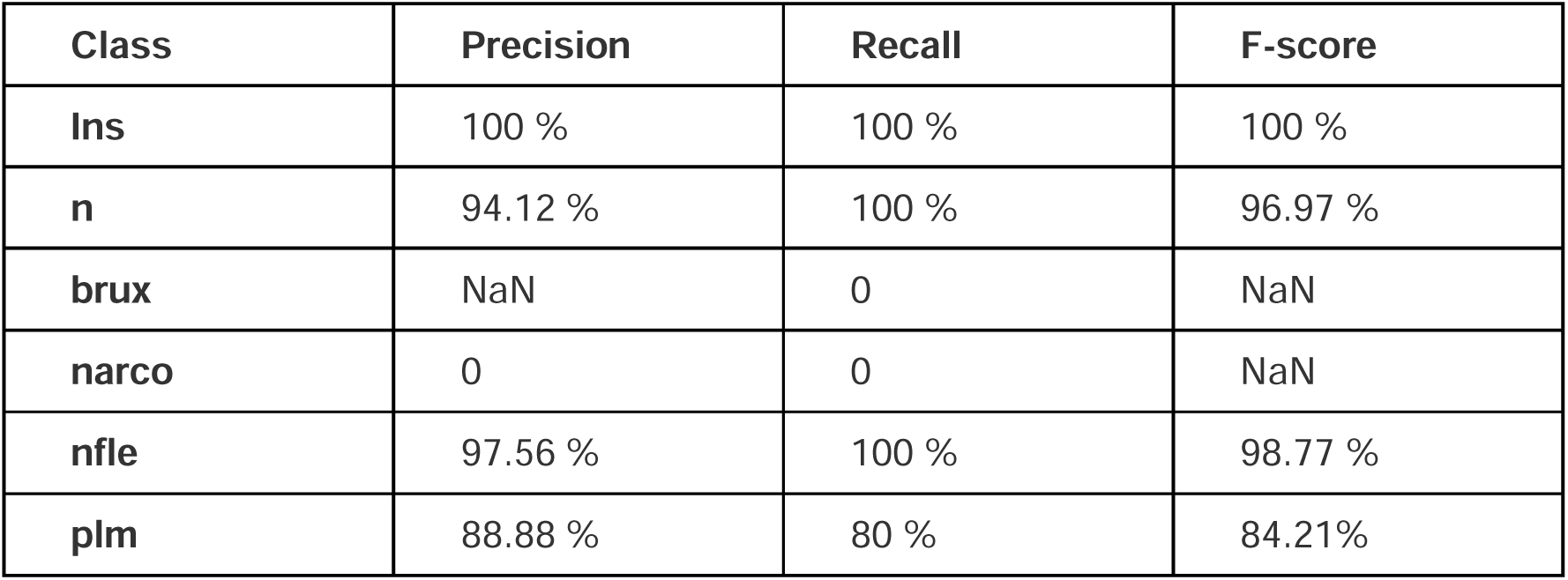
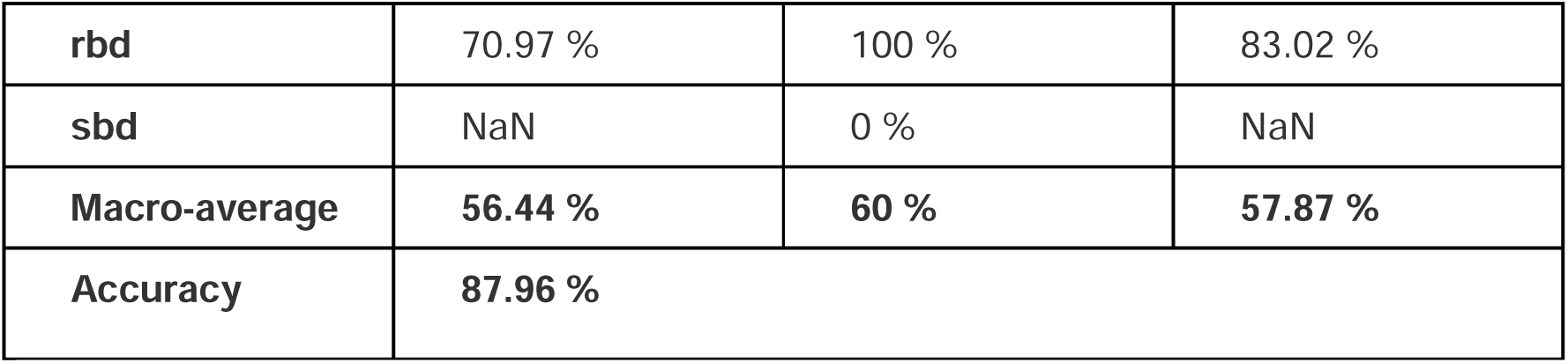
Precision, recall, F-score, and accuracy performance of the RF model for the eight-class experiment.

**Table 6.**
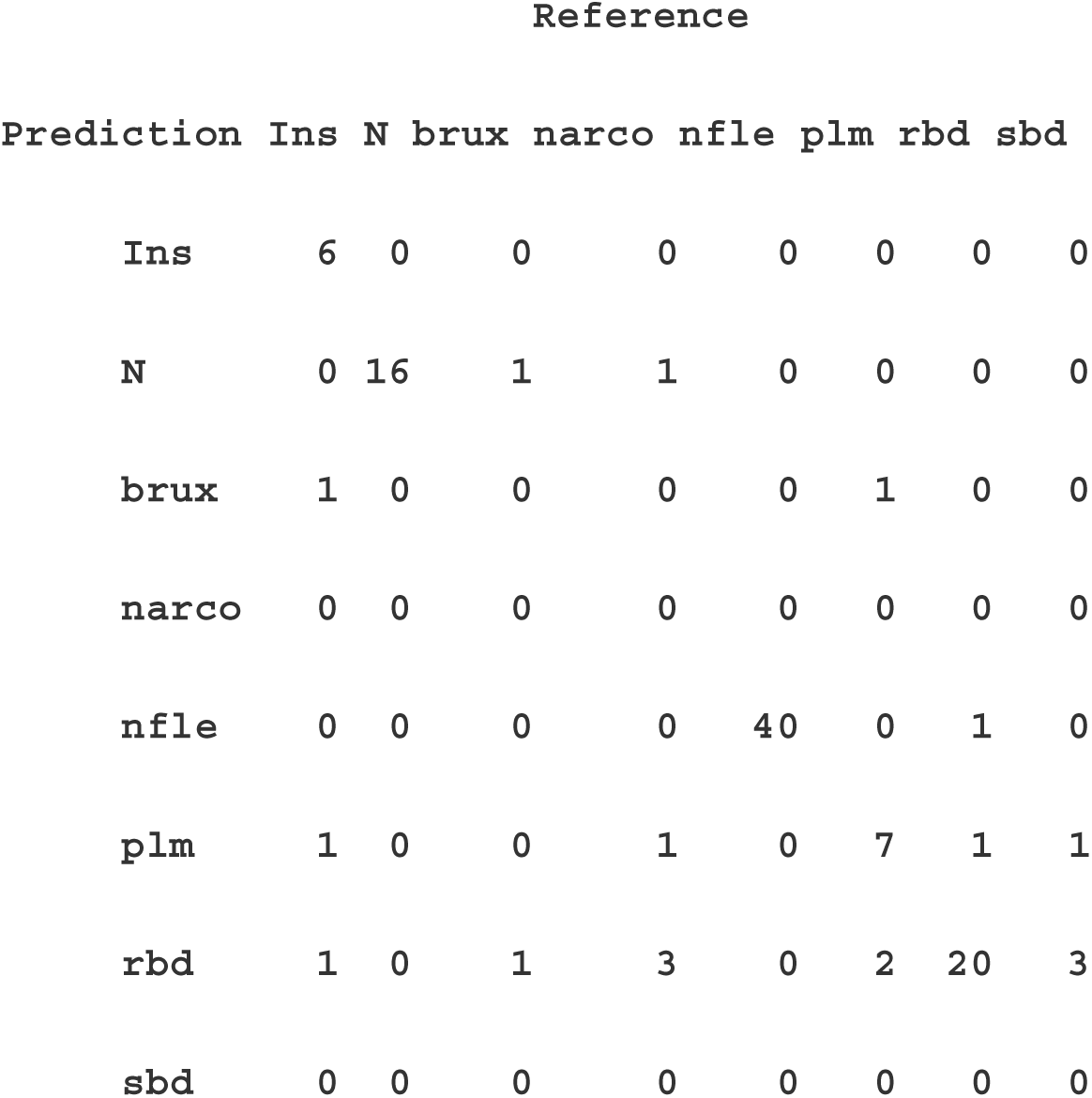
Confusion matrix of the RF model based on the selected features.

### 3.2 Performance of the RF model with the proposed SLEIS score on the Feature Set Level

Our RF model achieved a high performance of 96.74% in the seven-class scenario, with macro-average precision of 95.23%, macro-average recall of 90.71%, and macro-average *F*-score of 92.23% (**Table 7**). It is important to underline that the RF model misclassified two samples, one in rbd and one in sbd. The OOB estimate of error rate was 9.02%. **Table 8** summarizes the confusion matrix of the RF model for the eight-class approach. Based on the Gini Index and RF model, the four feature sets were ranked as follows: feature set 2 (CAP rates) = 25.42, feature set 3 (CAP cycles and sequences) = 14.62, feature set 1 (macrostructural features) = 13.14, and feature set 4 (CAP Phases A and B) = 12.90. Mean and standard deviation of the four features across the sleep stages are illustrated in **Fig.4**.

**Fig.4.**
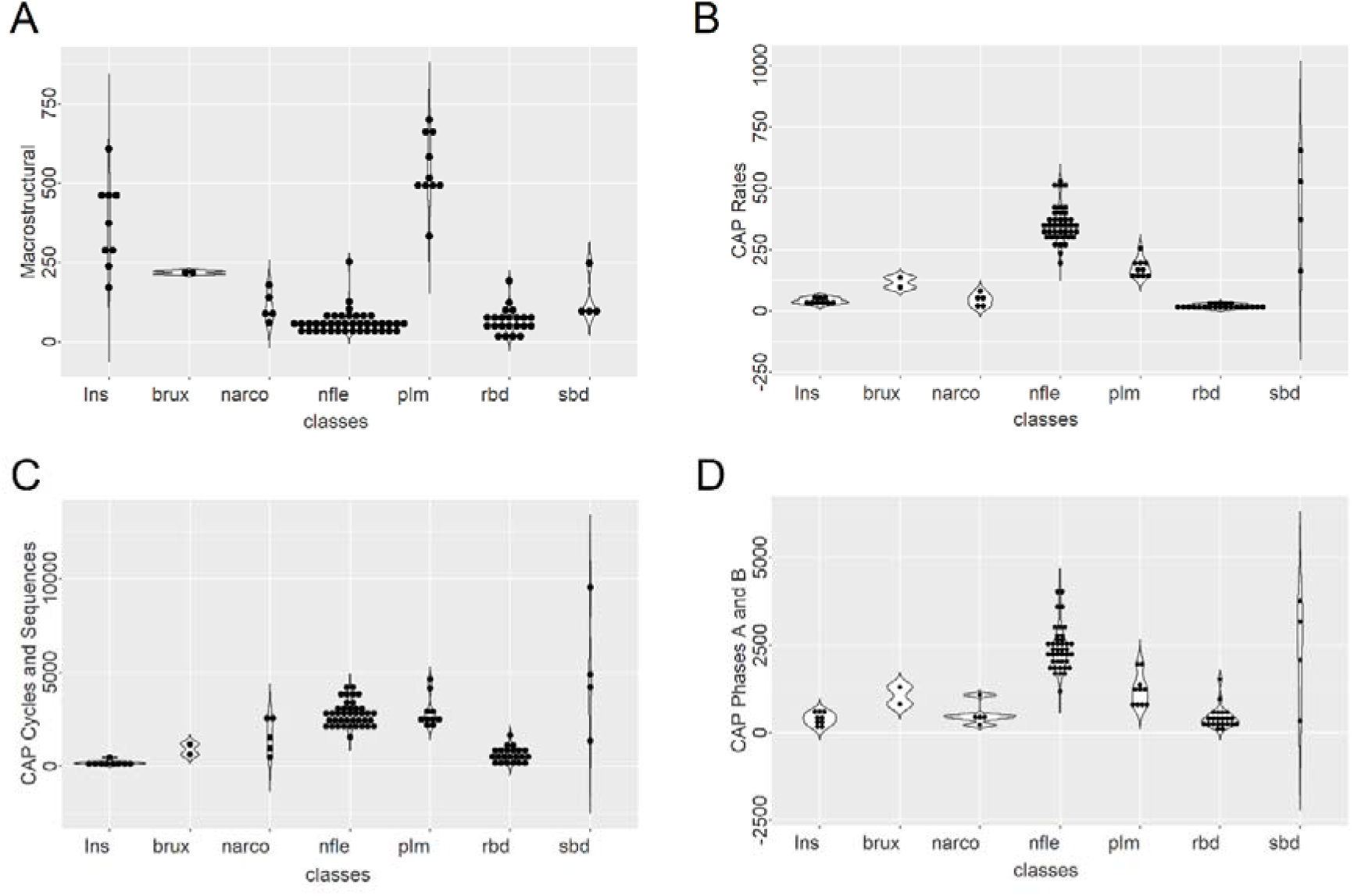
Mean and standard deviations of the four groups of features across the seven sleep disorders.

**Table 7.** Precision, recall, *F*-score, and accuracy performance of the RF model for the seven-class experiment based on the four feature sets.

| **Class** | **Precision** | **Recall** | **F-score** |
| --- | --- | --- | --- |
| **Ins** | 100 % | 100 % | 100 % |
| **brux** | 100 % | 100 % | 100 % |
| **narco** | 100 % | 60 % | 75 % |
| **nfle** | 100 % | 100 % | 100 % |
| **plm** | 100 % | 100 % | 100 % |
| **rbd** | 91.67 % | 100 % | 95.65 % |
| **sbd** | 75 % | 75 % | 75 % |
| **Macro-average** | **95.23 %** | **90.71%** | **92.23%** |
| **Accuracy** | **96.74%** |  |  |

**Table 8.** Confusion matrix of the RF model based on the four feature sets.

| Reference |  |  |  |  |  |  |  |
| --- | --- | --- | --- | --- | --- | --- | --- |
| Prediction | Ins | brux | narco | nfle | plm | rbd | sbd |
| Ins | 9 | 0 | 0 | 0 | 0 | 0 | 0 |
| brux | 0 | 2 | 0 | 0 | 0 | 0 | 0 |
| narco | 0 | 0 | 3 | 0 | 0 | 0 | 0 |
| nfle | 0 | 0 | 0 | 40 | 0 | 0 | 0 |
| plm | 0 | 0 | 0 | 0 | 10 | 0 | 0 |
| rbd | 0 | 0 | 1 | 0 | 0 | 22 | 1 |
| sbd | 0 | 0 | 1 | 0 | 0 | 0 | 3 |

### 3.3 Performance of the RF model with the proposed SLEIS score on the Feature Level

Our RF model achieved 100% performance in the seven-class scenario, with macro-average precision, recall, and F-score all at 100% (**Table 9**). The OOB estimate of error rate was 0%. **Table 10** summarizes the confusion matrix of the RF model for the seven-class approach. The nine most discriminative features, based on our RF model, with a Gini Index> 1 and a z-score > 1.5 across all features, are shown in **Fig. 5**.

**Fig.5.**
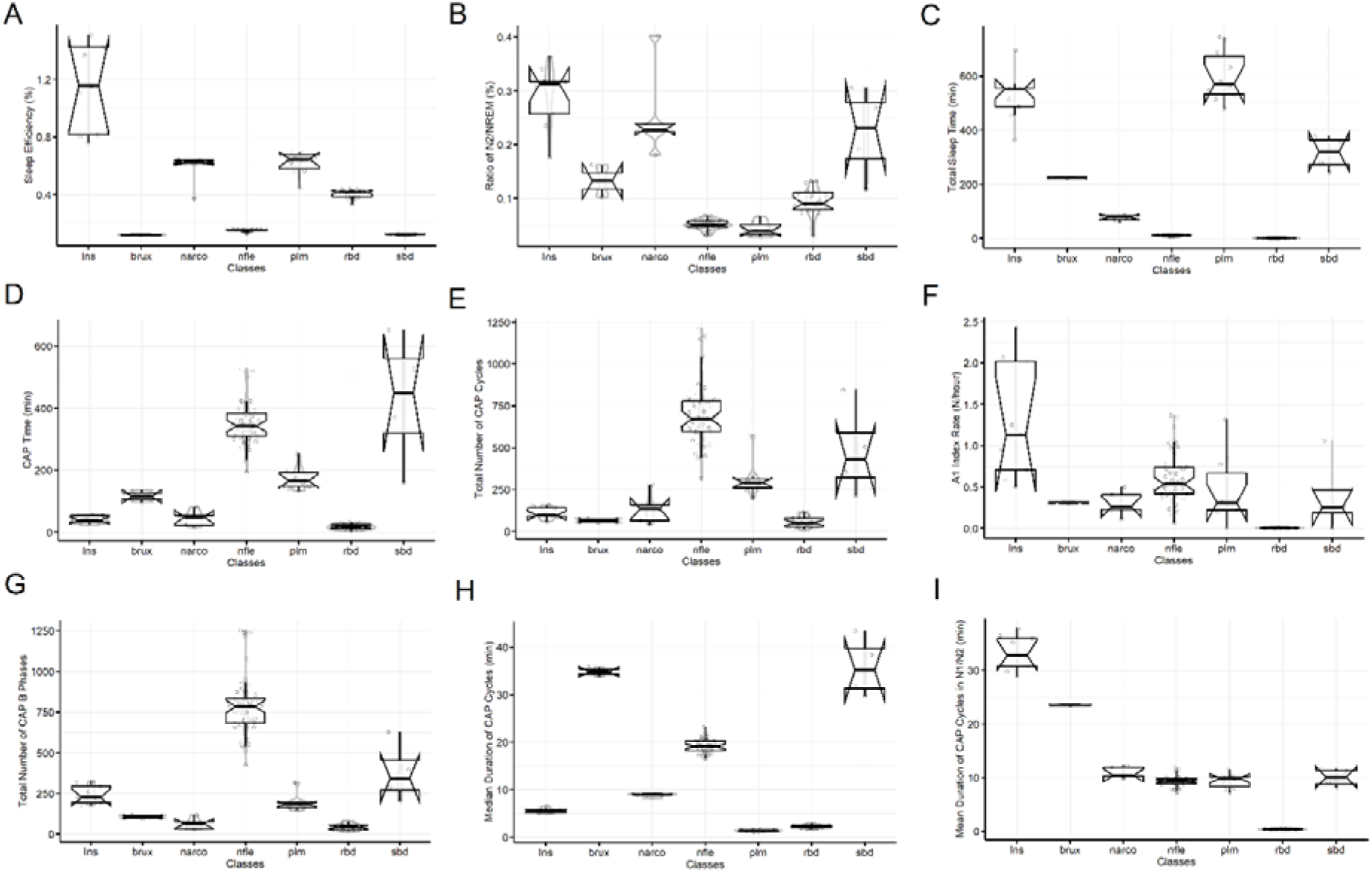
Mean and standard deviation of the nine most discriminative features for the seven sleep disorders.

**Table 9.** Precision, recall, *F*-score, and accuracy performance of the RF model for the seven-class experiment based on the ranked features.

| **Class** | **Precision** | **Recall** | **F-score** |
| --- | --- | --- | --- |
| **Ins** | 100 % | 100 % | 100 % |
| **brux** | 100 % | 100 % | 100 % |
| **narco** | 100 % | 100 % | 100 % |
| **nfle** | 100 % | 100 % | 100 % |
| **plm** | 100 % | 100 % | 100 % |
| **rbd** | 100 % | 100 % | 100 % |
| **sbd** | 100 % | 100 % | 100 % |
| **Macro-average** | **100 %** | **100 %** | **100 %** |
| **Accuracy** | **100 %** |  |  |

**Table 10.** Confusion matrix of the RF model based on the ranked features.

| Prediction | Ins | brux | narco | nfle | plm | rbd | sbd |
| --- | --- | --- | --- | --- | --- | --- | --- |
| Ins | 9 | 0 | 0 | 0 | 0 | 0 | 0 |
| brux | 0 | 2 | 0 | 0 | 0 | 0 | 0 |
| narco | 0 | 0 | 5 | 0 | 0 | 0 | 0 |
| nfle | 0 | 0 | 0 | 40 | 0 | 0 | 0 |
| plm | 0 | 0 | 0 | 0 | 10 | 0 | 0 |
| rbd | 0 | 0 | 0 | 0 | 0 | 22 | 0 |
| sbd | 0 | 0 | 0 | 0 | 0 | 0 | 4 |

## 4. Discussion

In the present study, we demonstrated the efficacy of a novel method to address intersleep-disorder variation in sleep macro- and microarchitecture by generating SLEIS scores that reflect an individual’s proclivity for sleep-disorder-related CAP alterations. This study, for the first time, extracted an exhaustive set of macrostructural and microstructural CAP sleep features from an open sleep disorder database. We determined the deviation of the sleep disorders groups from a healthy control group across all estimated features, quantifying the effect size using Cohen’s *d*. We generated two versions of the SLEIS score, representing a) the individual feature value multiplied by the corresponding Cohen’s *d*, and b) their cumulative weight over groups of features. Finally, we employed a Random Forest (RF) trained on original multi-features and weighted under the two versions of the SLEIS score to evaluate its performance in discriminating the seven sleep disorders. The performance of our RF model with the two SLEIS scores was successful (Macro-average precision: 92.23% / 100%), compared to the original unweighted features (Macro-average precision: 56.44%). To our knowledge, this is the first approach to use normative modeling weighting approaches for macro- and microstructural features, considering effect size as deviation from a normative sample. Moreover, for the first time, the present study assigns cumulative weights to individual features across different sleep disorders. Therefore, we presented and assessed a novel multi-feature marker of sleep instability, the SLEIS score, demonstrating its advantages over a conventional non-weighted approach.

Sleep is a dynamic and complex process regulated by mechanisms that guide alterations in NREM-REM sleep stages throughout the night. Sleep macro-architecture is characterized by the dominant slow wave sleep (SWS) (stage N3) in the first half of the night and the dominance of light sleep (N2) and REM in the second half of the night. Physiologically, this distinction in dominant sleep stages, including their distribution, is modulated by the homeostatic process and by systems that turn on/off REM (Brown et al. 2012). During the night, numerous transitions occur among these sleep stages, and brain activity alters in both amplitude and frequency over seconds and minutes. The detection of sleep stages (NREM-REM) from an EEG recording during the night is fundamental for constructing a hypnogram. The individual hypnogram supports the identification of CAPs, which are the main electrophysiological biomarker of sleep instability (Parrino et al. 2012).

CAP is a physiological EEG activity seen during NREM sleep. It consists of repeatable, spontaneous sequences with frequency or amplitude variations (phase A) that diverge from the background activity and recur at intervals of up to 60 seconds. Phase B follows phase A and is marked by a return to background activity. Both phases last between 2 and 60 seconds and together form a CAP cycle. Several CAP cycles form a CAP sequence, usually containing 5-6 cycles. A CAP sequence may begin and end in different sleep stages (Smerieri et al. 2007).

The absence of CAP for more than 60 seconds is scored as non-CAP (NCAP) and is considered a phase of stationary activity (without any phasic CAP-A event), regardless of the NREM sleep stage (Halász et al. 2014). Following this, based on the distribution of slow and fast EEG frequencies and the relevant phasic events of the microstructure of sleep (such as delta bursts and K-complexes), A phases are classified into three subtypes: A1, A2, and A3 (Terzano et al. 2001). Importantly, these phase A subtypes are not randomly distributed during the night; rather, their appearance is linked to the homeostatic, circadian, and ultradian mechanisms of sleep regulation (Parrino et al. 1993; Terzano et al. 2005; Halász et al. 2014).

Night sleep is characterized by several NREM-REM sleep cycles. Within every sleep cycle, we observe a deepening tendency from stage N1 to N4, followed by an ascending sequence of sleep stages from N3 toward N1, and finally REM sleep (Halász 2016). The CAP is considered a precursor of REM sleep; it contributes to the temporal sequence and alteration of NREM-REM sleep cycles, and, for these reasons, its metrics are essential for understanding sleep dynamics. Ιn the light of physiology, CAP A1 subtypes dominate the deepening branches of the first sleep cycles, and gradually decrease during the night, following the decline of the homeostatic processes. In contrast, CAP A2 and A3 predominate in the ascending branches of sleep cycles and can be considered precursors of REM sleep (Terzano et al. 2005). CAP A1 mostly consists of slow-wave activity (SWA), which is characterized by highly synchronized cortical neurons within the delta frequency range [0.5 - 4] Hz. SWA is elaborated during synaptic strength downscaling, which benefits network and cellular performance (Ferri et al. 2008; Cirelli and Tononi 2022).

The aforementioned description of sleep architecture and the relevant complex patterns of fluctuated activity suggest an underlying tuning to criticality (Lo et al. 2004; Assadzadeh and Robinson 2018; Wang et al. 2019; Lombardi et al. 2020). Neuronal avalanches are defined as cascades and bursts of neural activity that exhibit power-law distributions of size and duration. Therefore, neuronal avalanches can be recognized as a hallmark feature of critical dynamics in the sleep brain. A recent sleep study investigated the criticality of neuronal avalanches in normal human sleep and their relationship with sleep macro- and micro-architecture (Scarpetta et al. 2023). The authors demonstrated that avalanche dynamics are modulated by NREM-REM sleep cycles and that scale-free neuronal avalanche occurrence during NREM sleep stages correlates with CAP activation phases. The aforementioned work demonstrated a potential functional link between CAP phases and brain tuning to criticality. In detail, neuronal avalanches’ occurrence: a) increases in N2-N4 sleep stages during sleep deepening periods, b) decreases during the ascending branch of sleep from N4 toward stage N1, and c) correlates with the occurrence of CAP phase A, with stronger/weaker positive correlations with CAP A1 subtypes/CAP A2, A3 subtypes. SWA and the appearance of CAP-A1 phases play important roles in sleep-dependent learning processes (Ferri et al. 2008). For that reason, a functional link between avalanche dynamics and sleep-dependent cognitive/learning processes seems logical (Scarpetta et al. 2023).

On the one hand, CAP can reflect the sleeping brain’s reaction to any endogenous or exogenous disturbance. On the other hand, the CAP A phase has been interpreted as a kind of gate through which pathological events are more likely to occur (Parrino et al. 2006). CAP is a physiological component of NREM sleep observed in normal individuals (Terzano et al. 1985) and shows age-dependent features across the lifespan (Parrino et al. 1998). Increased amounts of CAP are often observed in sleep-disordered breathing (SDB), as well as in insomnia (Terzano et al. 2003), in sleep movement disorders (periodic leg movements (PLM) (Parrino et al. 1996), restless leg syndrome (RLS) (Hening 2004)), in bruxism ((Zucconi and Ferini-Strambi 2000; Kato et al. 2003), in parasomnias like REM behavior disorder (RBD), and in epileptic diseases such as nocturnal frontal lobe epilepsy (NFLE) (Zucconi and Ferini-Strambi 2000; Kato et al. 2003). Abnormal amounts of CAP can also be found in hypersomnias such as narcolepsy (Terzano et al. 2006; Poryazova et al. 2011).

In normal sleepers, arousals are more frequent during CAP (40/hour) than during total sleep time (18/hour), NREM sleep (20/hour), or REM sleep (12/hour) (Terzano et al. 2002). CAP not only attracts arousals, but also organizes the distribution of K-complexes and delta bursts in NREM sleep. When arousals overlap with CAP phase subtypes A2 or A3, the CAP rules allow a clearer evaluation of sleep instability, marked by delta bursts, K-complexes, and CAP phase subtypes A1. Thus, focusing only on arousals neglects other important cortical-subcortical phenomena in this complex structural pattern (Parrino et al. 2012; Scarpetta et al. 2023).

The locus coeruleus (LC), a small nucleus in the brainstem, serves as the primary source of noradrenaline (NA, also known as norepinephrine—a neurotransmitter involved in attention and arousal) for the entire central nervous system. The LC can precisely neuromodulate distinct brain regions and functions—including attention, arousal, and stress responses—over specific time periods (Breton-Provencher et al. 2021). As the main component of the ascending arousal system, the LC functions in parallel with other cholinergic and monoaminergic nuclei, which are groups of nerve cells that release acetylcholine or other monoamines, to regulate sleep stages and wakefulness (Saper et al. 2010). During non-rapid eye movement (NREM) sleep, the LC exhibits infra-slow oscillatory activity (very slow, rhythmic fluctuations in neuronal activity, ranging from 0.01 to 1 Hz) with a periodicity of around 30–50 seconds (Kjaerby et al. 2020). The timing of these oscillations aligns with the timescale of the cyclic alternating pattern (CAP), a type of EEG pattern in sleep, suggesting a potential functional interaction between CAP dynamics and LC-driven noradrenergic rhythms (Kjaerby et al. 2020).

In the present study, we attempted to identify a multi-feature marker of sleep instability, focusing on CAP sequences and complementary macrostructural features. We aimed to define and evaluate a common sleep instability marker across seven sleep disorders by using a healthy control group as a reference. For that purpose, we considered the effect size of each feature size relative to a normative sample, and we finally weighted individual features across various sleep disorders, both individually and cumulatively within groups of features. Therefore, we presented and evaluated a novel multi-feature marker of sleep instability, the SLEIS score, above a conventional non-weighted approach. Our RF model, in conjunction with both versions of SLEIS, outperformed non-weighted features and achieved an absolute performance in the individual approach (100%).

Our analysis focused on investigating microstructural and macrostructural features, both qualitatively and quantitatively, to discriminate sleep disorders from healthy controls and between them. SLEIS score can be considered as a general sleep instability index that is simultaneously sensitive to various sleep disorders. It can also be applied as a neurophysiological marker of disorders of arousal in parasomnias and sleep disorders (e.g., NFLE, RBD), complementing current knowledge of macro- and microstructural alterations (Bergmann et al. 2024).

It is of paramount importance to investigate the dynamics relevant to CAP function, as well as the relationships among neuronal avalanches, sleep architecture, and self-organization toward criticality (Scarpetta et al. 2023). Complementary, we should explore the EEG frequency components of CAP, like the distribution of frequencies across the scalp and virtual sources, and their dominant frequency band in the infraslow activity (Onton et al. 2016). CAP A phases, as previously noted, exhibit distinct frequency ranges: A1 (0.25–2.5 Hz) and A3 (7–12 Hz). Source localization using low-resolution brain electromagnetic tomography (LORETA) indicates that A1 generators are mainly in the frontal midline cortex, whereas A3 generators are distributed across midline and hemispheric parietal and occipital regions. The structurally coordinated association between two distinct CAP frequency components located in anatomically distant brain areas indicates functional intercoupling between them. This functional intercoupling could be altered in sleep disorders.

### Limitations

The first limitation of the present study concerns the number of samples per sleep disorder and the underrepresentation of certain sleep disorder classes. Secondly, our analysis focused on annotated epochs for both sleep stages (W/N1/N2/N3/N4/REM) and CAP phases (A1/A2/A3/B), as well as the time ranges of CAP cycles and sequences. To take advantage of this work, automatic CAP Phases A and B classification is important (Mariani et al. 2010, 2013; Ferri et al. 2012; Dhok et al. 2020; Tramonti Fantozzi et al. 2021; Kahana et al. 2023; Sharma et al. 2023; Telangore et al. 2025). Automatic detection of the start-end time sample point, along with automatic sleep stage classification in healthy individuals (Dimitriadis et al. 2018b; Jain and G 2024; Chen et al. 2025) and in sleep disorders (Sharma et al. 2021, 2022), is also crucial. Manual annotation of sleep stages and CAP phases by experts is time-consuming. For that reason, the clinical applications of CAP analysis remain limited. Thirdly, due to limitations in the CAP sleep study, we could not correlate the extracted features with clinical variables across the sleep disorders.

### Conclusions

In this study, we used the healthy control group from this unique database as a common reference for seven sleep disorders, a first, to our knowledge. This provided robust prior effect size weights and helped parse heterogeneity in subjects and disorders by accounting for individual macrostructural-microstructural profiles, improving disorder-specific sensitivity. The trained RF model’s absolute performance with proposed SLEIS scores, compared to original feature weights, validated our hypothesis. Using a larger normative lifespan database as a reference is a promising research direction. Further, investigating how EEG features—such as power spectra and connectivity—relate to the extracted features here, including SLEIS markers, will help reveal the neurophysiology of sleep disorders. Systematic comparison of macrostructural, microstructural, and SLEIS markers with clinical diagnostic features is warranted. Various automated methods for sleep stage classification and for detecting CAP phase start and end times will support SLEIS marker computation.

## Author contributions

**Stavros I Dimitriadis**: conceptualization, supervision, data curation, methodology, formal analysis, software, writing (original draft), and performed the machine learning analysis.

**Christos Salis**: data curation, formal analysis, software, performed the feature extraction analysis, writing (reviewing and editing).

## Funding

This research has been conducted in the absence of any funding support.

## Conflict of interest

The authors declare that the research was conducted in the absence of any commercial or financial relationships that could be construed as a potential conflict of interest.

## Data availability statement

The data supporting the findings of the present study are openly available at the following URL: https://physionet.org/content/capslpdb/1.0.0/.

The original features and the SLEIS scores will be released in our github’s website upon acceptance of the manuscript.

https://github.com/stdimitr/CAP_Sleep_Disorders/tree/main

## Acknowledgments

We would like to acknowledge researchers, staff, and technicians who made this dataset available to the public. Special thanks also to the staff and scientists who keep the Physionet database active.

## Abbreviations

NREM: non-rapid eye movement
REM: rapid eye movement
SWS: slow wave sleep
SWA: slow wave activity
SCN: suprachiasmatic nucleus
LC: locus coeruleus
NA: noradrenaline
CAP: cyclic alternating pattern
NCAP: non-CAP
CAPs: CAP sequences
EEG: electroencephalography
LORETA: low resolution brain electromagnetic tomography
RF: Random Forest
OOB: out-of-bag (OOB) error
SLEIS: sleep instability score

## Notes

### Competing Interest Statement

The authors have declared no competing interest.

### Funding Statement

This study did not receive any funding

### Author Declarations

The study used (or will use) ONLY openly available human data that were originally available at the following website: following URL: https://physionet.org/content/capslpdb/1.0.0/.

### Summary of Updates

I mistakenly wrote in the title 'Acyclic' instead of 'Alternating'.

